# Overweight and Obesity as Predictors of Post-acute Sequelae of SARS-Cov-2 Infection: Findings from the RECOVER Initiative

**DOI:** 10.1101/2024.06.12.24308868

**Authors:** Ting Zhou, Bingyu Zhang, Dazheng Zhang, Qiong Wu, Michael J. Becich, Saul Blecker, Jiajie Chen, Nymisha Chilukuri, Elizabeth A. Chrischilles, Haitao Chu, Leonor Corsino, Carol R. Geary, Mady Hornig, Susan Kim, David M. Liebovitz, Vitaly Lorman, Yiwen Lu, Chongliang Luo, Hiroki Morizono, Abu SM. Mosa, Nathan M. Pajor, Suchitra Rao, Hanieh Razzaghi, Srinivasan Suresh, Yacob G. Tedla, Leah Vance Utset, Youfa Wang, David A. Williams, Margot Gage Witvliet, Caren Mangarelli, Ravi Jhaveri, Christopher B. Forrest, Yong Chen

## Abstract

**IMPORTANCE:** Obesity increases the severe COVID-19 risk. Whether obesity is associated with an increased risk of post-acute sequelae of SARS-Cov-2 infection (PASC) among pediatrics, independent of its impacts on acute infection severity, is unclear.

**OBJECTIVE:** To quantify the association between body mass index (BMI) status before SARS-CoV-2 infection and pediatric PASC risk, controlling for acute infection severity.

**DESIGN:** Retrospective cohort study occurred from March 2020 to May 2023, with a minimal follow-up of 179 days.

**SETTING:** Twenty-six US children’s hospitals.

**PARTICIPANTS:** Individuals aged 5-20 years with SARS-CoV-2 infection.

**EXPOSURES:** Elevated BMI status assessed before infection.

**MAIN OUTCOMES AND MEASURES:** To identify PASC, we first used the ICD-10-CM code specific for post-COVID-19 conditions, and a second approach used clusters of symptoms and conditions that constitute the PASC phenotype. BMI was assessed within 18 months before infection; the measure closest to the index date was selected. Relative risk (RR) for BMI-PASC association was quantified by Poisson regression models, adjusting for sociodemographic, acute COVID severity, and other clinical factors.

**RESULTS:** Among the 172136 participants included, the median age of BMI assessment and cohort entry were 12.8 and 13.2 years, 1402 (0.8%) were identified as having PASC with the ICD-10-CM code, and 74317 (43.2%) had ≥1 incident occurrence of PASC symptoms and conditions. Compared with participants with a healthy weight, those who had overweight, obesity, and severe obesity had 4.7% (RR, 1.047; 95% CI, 0.868-1.263), 25.4% (RR, 1.254; 95% CI, 1.064-1.478) and 42.1% (RR, 1.421; 95% CI, 1.253-1.611) higher risk of PASC when identified using the diagnosis code, respectively. The risk for any occurrences of PASC symptoms and conditions also increased in overweight (RR, 1.030; 95% CI, 0.982-1.080), obesity (RR, 1.108; 95% CI, 1.064-1.154), and severe obesity (RR, 1.174; 95% CI, 1.138-1.213), and that for total incident occurrences increased, too, in overweight (RR, 1.053; 95% CI, 1.000-1.109), obesity (RR, 1.137; 95% CI, 1.088-1.188), and severe obesity (RR, 1.182; 95% CI, 1.142-1.223).

**CONCLUSIONS AND RELEVANCE:** Elevated BMI was associated with a significantly increased PASC risk in a dose-dependent manner. The biological mechanisms for this association should be investigated in future research.

**Key Points:** *Question:* Do children, adolescents, and young adults with overweight and obesity have increased risk of developing post-acute sequelae of SARS-Cov-2 infection (PASC)?

*Findings:* Overweight, obesity, and severe obesity were associated with significantly increased risk of pediatric PASC. Compared with pediatrics with body mass index in the healthy range, those who were overweight, obesity, or severely obesity had an increased incidence of 4.7%, 25.4%, and 42.1% of PASC, respectively.

*Meaning:* Overweight and obesity are important risk factors for pediatric PASC. The biological mechanisms for this association should be investigated in the future research.

---

Post-acute sequelae of SARS-CoV-2 infection (PASC) encompasses a broad and heterogeneous array of persistent, relapsing, or newly emerging symptoms persisting beyond at least 4 weeks after the acute phase of COVID-19.^1–4^ This condition exhibits multifaceted involvement across various organ systems.^5–8^ The prevalence of pediatric PASC with SARS-CoV-2 infection varies across studies with reported rates ranging from 1.6% to 70%.^9–12^ PASC continues to pose a significant threat to children and there is an urgent imperative for a deeper understanding of the pediatric PASC causes, which is a research priority underscored in the updated National Institute for Health and Care Excellence guideline.^13^ This imperative persists, emphasizing the ongoing need to unravel the complexities surrounding pediatric PASC even after the pandemic.

Obesity is now one of the most common chronic diseases in the US, impacting over 40% of adults and about 20% of children.^14,15^ The association of obesity with severe adverse outcomes was seen again with the onset of the COVID-19 pandemic.^16^ The association between obesity as a risk factor and PASC has been widely discussed, with a predominant focus on adults.^17,18^ Some studies have reported that overweight or obesity is associated with an elevated risk of PASC^18,19^ across different timelines, irrespective of whether the definition of PASC extends to 12 weeks^18^ or 4 months.^19^ However, the nuances of this relationship become apparent when considering the specific timeline for defining PASC. For example, Sudre et al. observed that increasing body mass index (BMI) and obesity were linked to higher odds of PASC lasting for more than 4 weeks, but this association did not extend to PASC lasting for more than 12 weeks.^20^

The major concern is the limited attention given to the pediatric population, as the previous investigations predominantly concentrated on adults. There is hardly any discussion regarding the pediatric BMI-PASC association, which underscores a crucial research gap. To address these limitations and enhance our understanding, large-scale studies using routinely available healthcare data among pediatric population are needed. Our study conducted an extensive analysis utilizing EHR data sourced from 26 US children’s hospitals. The primary objective was to explore the relationship between BMI status before SARS-CoV-2 infection and a range of PASC definitions among both hospitalized and non-hospitalized pediatric populations with COVID-19 infection, controlling for acute infection severity. We also assessed associations between BMI status and the number of PASC symptoms and conditions accounting for sociodemographic and clinical risk factors. Importantly, the insights obtained will play an influential role in informing preventive and clinical management strategies, assisting healthcare providers in identifying at-risk pediatric patients to develop PASC.

## Methods

### Data Sources

This study is part of the National Institutes of Health Researching COVID-19 to Enhance Recovery (https://recovercovid.org/), which aims to understand, treat, and prevent PASC. Twenty-six institutions contributed to the data (see eTable 1 in the Supplement for details).

We conducted a retrospective cohort study from March 2020 to May 2023 by including documented SARS-CoV-2 infected patients under the age of 21 who had at least one visit within the baseline period of 18 months to 7 days prior to the index date and had at least one visit within the follow-up period of 28 days to 179 days after the index date. Documented SARS-CoV-2 infections were defined by SARS-CoV-2 polymerase chain reaction, antigen or serology positive, or diagnosis of COVID-19, PASC or multisystem inflammatory syndrome (MIS). The index date was set as either the earliest date of positive tests or COVID-19 diagnoses or 28 days before PASC/MIS diagnosis.

Participants were excluded if they aged below 5 at the time of assessing BMI due to their potential for more dramatic BMI variations during the baseline period, and if they had genetic syndromes associated with obesity or any conditions signaling a need for weight gain or a medical cause of altered weight tendencies during the baseline period (eTable 2 in the Supplement). The participants selection process is summarized in eFigure 1 in the Supplement.

### Defining BMI Status

When multiple BMI (weight (kg)/height (m)^2^) measures were available during the baseline period, we selected the measure closest to the index date. For participants aged 5-19 years, according to the age-sex– specific BMI percentiles based on CDC Growth Charts, the BMI status was categorized into healthy weight (5^th^ to less than 85^th^ percentile), overweight (85^th^ to less than 95^th^ percentile), obesity (95^th^ percentile to less than 120% of the 95^th^ percentile), and severe obesity (120% of the 95^th^ percentile or greater );^21^ for participants aged above 19 years, these four categories were divided by BMI of 18.5 to less than 25, 25 to less than 30, 30 to less than 40, and 40 or more within the baseline period.^22^

### Defining PASC

The outcome was assessed within the follow-up period. We first used ICD-10-CM codes, U09.9, specific for post-COVID-19 condition to indicate PASC. For this, we created a binary outcome, PASC (U09.9), which was defined as positive if anyone who was identified as having PASC with a U09.9 diagnosis code.

Second, we used clusters of symptoms and health conditions previously shown to constitute the PASC phenotype based on experts’ suggestions,^23, 24^ including abdominal pain, abnormal liver enzyme, acute kidney injury, acute respiratory distress syndrome, arrhythmias, cardiovascular signs and symptoms, changes in taste and smell, chest pain, cognitive dysfunction, fatigue and malaise, fever and chills, fluid and electrolyte imbalances, generalized pain, hair loss, headache, heart disease, mental health disorders, musculoskeletal pain, myocarditis, myositis, postural orthostatic tachycardia syndrome or dysautonomia, respiratory signs and symptoms, skin symptoms, thrombophlebitis and thromboembolism. We assessed the incident occurrences of these 24 PASC symptoms and conditions within the follow-up period, but that did not occur during the baseline period, and then a binary and a count outcome were created. The binary outcome is defined as positive if any of these conditions occurred, and the count outcome is defined as the total incident occurrences of these conditions.

### Covariates

Age at BMI status assessment and age at cohort entry were collected. Other demographic characteristics included sex (male/female), and race and ethnicity (Asian American/Pacific Islander, Hispanic, Non-Hispanic Black, and Non-Hispanic White). The predominating COVID-19 virus variant was categorized as pre-Alpha (2020-03-01∼ 2021-03-31), Alpha (2021-04-01∼2021-06-30), Delta (2021-07-01∼2021-12-31), and Omicron (2022-01-01∼2022-12-01).^25^ We used numbers of emergency department/ outpatient department/inpatient department visits, medications or prescriptions, and negative COVID-19 tests up to 24 months before index date as healthcare utilization metrics, and the Pediatric Medical Complexity Algorithm (PMCA)^26^ index to address levels of medical complexity comorbidity (no/non-complex/complex chronic condition) for the pediatrics. We also classified participants based on the acute COVID-19 severity as asymptomatic, mild, moderate, and severe.^27^ Doses of COVID-19 vaccine prior to infection and interval since last COVID-19 vaccination date (no vaccine/<4 months/≥4 months), and type of insurance (private/public/others) were also considered in the secondary analyses.

### Statistical Analysis

We first presented the demographic and clinical characteristics across different BMI statuses. Comparisons of PASC (U09.9) and potential PASC symptoms and conditions were then made by BMI status. For the count outcome, total incident occurrences of PASC symptoms and conditions, we used Poisson regression to assess its association with BMI status by estimating the relative risk (RR) and 95% confidence interval (CI), adjusting for demographic characteristics (age, sex, race/ethnicity), predominant variant, healthcare utilization metrics prior to cohort entry, PMCA index, and acute COVID-19. For binary outcomes, PASC (U09.9) and any incident occurrences of PASC symptoms and conditions, modified Poisson regression^28^ was employed, accounting for possible variance overestimation and adjusting for the factors described above. Analysis of variance was used for the linear trend test. Subgroup analyses were performed by age (<18 or ≥18 years) and PMCA index (no chronic condition or non-complex/complex chronic condition), and by race/ethnicity as disparities exist in the BMI status among children and the severity of COVID-19 by race/ethnicity to test for potential effect modification.

We performed sensitivity analyses to validate our findings. First, we excluded participants whose BMI status assessment date was beyond 6 months before cohort entry as pediatric BMI tended to increase during the pandemic.^29^ Second, participants whose entrance date was before the time U09.9 was released (i.e.. October 1, 2021) were excluded considering possible low usage of U09.9 by clinicians when the code was first introduced. We then excluded patient confirmed by serology test after November 2022 due to the test’s possible accuracy issues. Fourth, to investigate whether the observed associations were explained by the acute COVID-19 severity, we excluded participants with moderate or severe infection. Fifth, as vaccination against COVID-19 may reduce PASC risk,^30,31^ we additionally adjusted for vaccination status before infection. Sixth, insurance type was further adjusted to examine whether socioeconomic factors might account for the association. Seventh, participants with diabetes were excluded as diabetes has been reported as a risk factor for PASC.^32,33^ Eighth, we excluded obese participants without diabetes using weight-loss drugs during the baseline period to investigate whether weight-loss drugs impact the association. Nineth, we conducted an analysis based on primary care sites to ascertain the generalizability of our findings within a community-based primary care setting. We also used foreign body in ear as a negative control outcome to account for the impacts due to residual bias.^34,35^ All analyses were conducted using R version 4.1.2 (The R Foundation).

## Results

After excluding participants who were underweight (n=221) and missed BMI status (n=67451), a total of 172136 participants were included in the analysis. Participants missing BMI status data were more prone to be younger, male, Non-Hispanic White and Non-Hispanic Black, be infected after the pre-Alpha wave, have lower complex chronic conditions, use healthcare utilization less, and be less likely to develop PASC (eTable 3 in the Supplement). The median time from BMI status assessment to COVID-19 infection was 4.1 months (IQR 1.6-8.6). Of all the participants, 90187 (52.4%) were female, 87275 (50.7%) were Non-Hispanic White, and 85613 (49.7%) had obesity or severe obesity (Table 1).

**Table 1.**
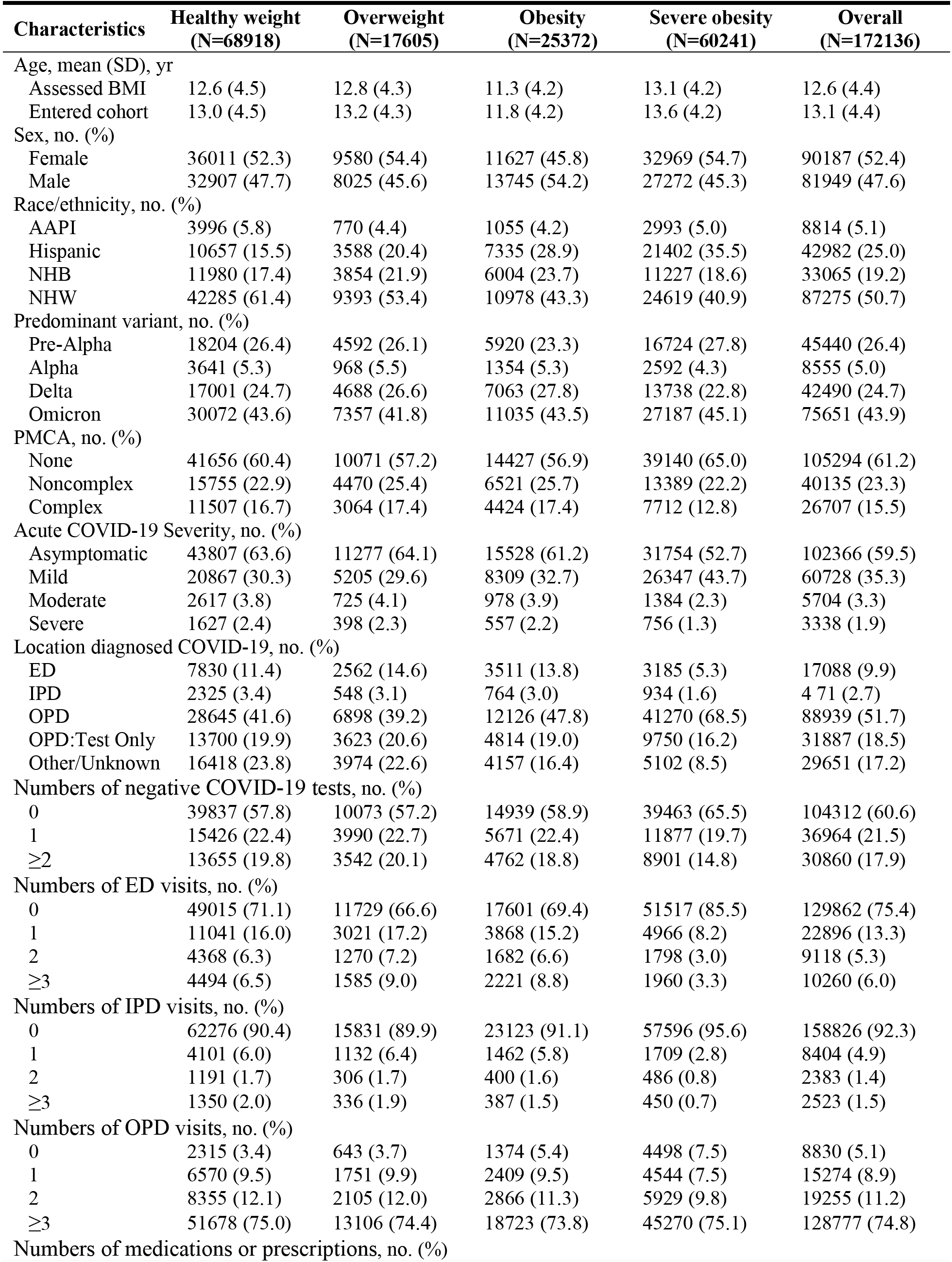
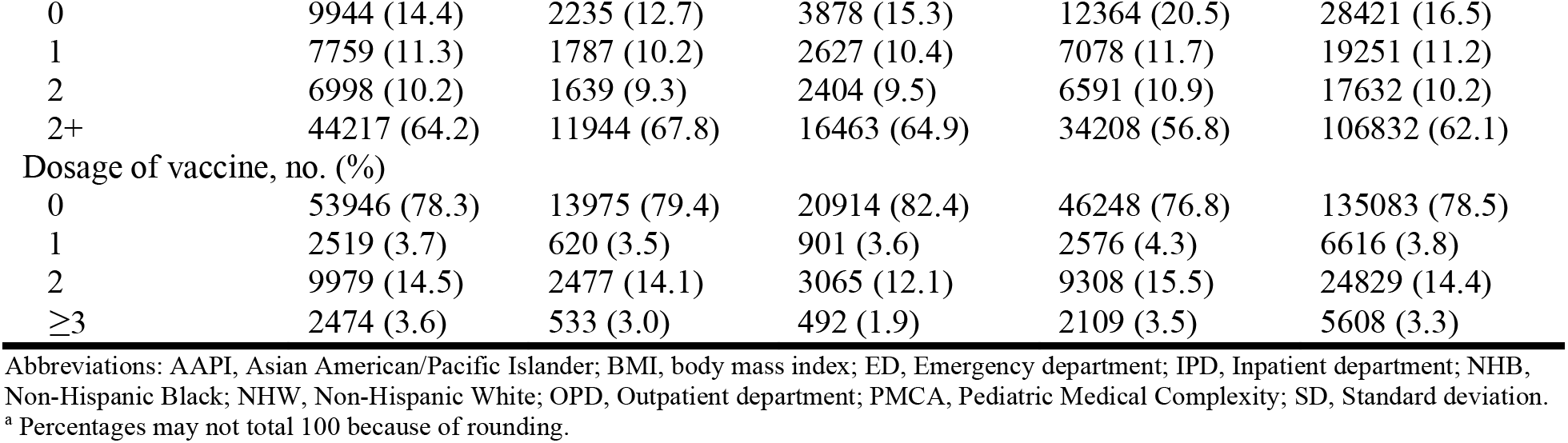
Characteristics by BMI status prior to the SARS-CoV-2 infection pediatric participants ^a^.

| Characteristics | Healthy weight<br>(N=68918) | Overweight<br>(N=17605) | Obesity<br>(N=25372) | Severe obesity<br>(N=60241) | Overall<br>(N=172136) |
| --- | --- | --- | --- | --- | --- |
| Age, mean (SD), yr |  |  |  |  |  |
| Assessed BMI | 12.6 (4.5) | 12.8 (4.3) | 11.3 (4.2) | 13.1 (4.2) | 12.6 (4.4) |
| Entered cohort | 13.0 (4.5) | 13.2 (4.3) | 11.8 (4.2) | 13.6 (4.2) | 13.1 (4.4) |
| Sex, no. (%) |  |  |  |  |  |
| Female | 36011 (52.3) | 9580 (54.4) | 11627 (45.8) | 32969 (54.7) | 90187 (52.4) |
| Male | 32907 (47.7) | 8025 (45.6) | 13745 (54.2) | 27272 (45.3) | 81949 (47.6) |
| Race/ethnicity, no. (%) |  |  |  |  |  |
| AAPI | 3996 (5.8) | 770 (4.4) | 1055 (4.2) | 2993 (5.0) | 8814 (5.1) |
| Hispanic | 10657 (15.5) | 3588 (20.4) | 7335 (28.9) | 21402 (35.5) | 42982 (25.0) |
| NHB | 11980 (17.4) | 3854 (21.9) | 6004 (23.7) | 11227 (18.6) | 33065 (19.2) |
| NHW | 42285 (61.4) | 9393 (53.4) | 10978 (43.3) | 24619 (40.9) | 87275 (50.7) |
| Predominant variant, no. (%) |  |  |  |  |  |
| Pre-Alpha | 18204 (26.4) | 4592 (26.1) | 5920 (23.3) | 16724 (27.8) | 45440 (26.4) |
| Alpha | 3641 (5.3) | 968 (5.5) | 1354 (5.3) | 2592 (4.3) | 8555 (5.0) |
| Delta | 17001 (24.7) | 4688 (26.6) | 7063 (27.8) | 13738 (22.8) | 42490 (24.7) |
| Omicron | 30072 (43.6) | 7357 (41.8) | 11035 (43.5) | 27187 (45.1) | 75651 (43.9) |
| PMCA, no. (%) |  |  |  |  |  |
| None | 41656 (60.4) | 10071 (57.2) | 14427 (56.9) | 39140 (65.0) | 105294 (61.2) |
| Noncomplex | 15755 (22.9) | 4470 (25.4) | 6521 (25.7) | 13389 (22.2) | 40135 (23.3) |
| Complex | 11507 (16.7) | 3064 (17.4) | 4424 (17.4) | 7712 (12.8) | 26707 (15.5) |
| Acute COVID-19 Severity, no. (%) |  |  |  |  |  |
| Asymptomatic | 43807 (63.6) | 11277 (64.1) | 15528 (61.2) | 31754 (52.7) | 102366 (59.5) |
| Mild | 20867 (30.3) | 5205 (29.6) | 8309 (32.7) | 26347 (43.7) | 60728 (35.3) |
| Moderate | 2617 (3.8) | 725 (4.1) | 978 (3.9) | 1384 (2.3) | 5704 (3.3) |
| Severe | 1627 (2.4) | 398 (2.3) | 557 (2.2) | 756 (1.3) | 3338 (1.9) |
| Location diagnosed COVID-19, no. (%) |  |  |  |  |  |
| ED | 7830 (11.4) | 2562 (14.6) | 3511 (13.8) | 3185 (5.3) | 17088 (9.9) |
| IPD | 2325 (3.4) | 548 (3.1) | 764 (3.0) | 934 (1.6) | 4 71 (2.7) |
| OPD | 28645 (41.6) | 6898 (39.2) | 12126 (47.8) | 41270 (68.5) | 88939 (51.7) |
| OPD:Test Only | 13700 (19.9) | 3623 (20.6) | 4814 (19.0) | 9750 (16.2) | 31887 (18.5) |
| Other/Unknown | 16418 (23.8) | 3974 (22.6) | 4157 (16.4) | 5102 (8.5) | 29651 (17.2) |
| Numbers of negative COVID-19 tests, no. (%) |  |  |  |  |  |
| 0 | 39837 (57.8) | 10073 (57.2) | 14939 (58.9) | 39463 (65.5) | 104312 (60.6) |
| 1 | 15426 (22.4) | 3990 (22.7) | 5671 (22.4) | 11877 (19.7) | 36964 (21.5) |
| ≥2 | 13655 (19.8) | 3542 (20.1) | 4762 (18.8) | 8901 (14.8) | 30860 (17.9) |
| Numbers of ED visits, no. (%) |  |  |  |  |  |
| 0 | 49015 (71.1) | 11729 (66.6) | 17601 (69.4) | 51517 (85.5) | 129862 (75.4) |
| 1 | 11041 (16.0) | 3021 (17.2) | 3868 (15.2) | 4966 (8.2) | 22896 (13.3) |
| 2 | 4368 (6.3) | 1270 (7.2) | 1682 (6.6) | 1798 (3.0) | 9118 (5.3) |
| ≥3 | 4494 (6.5) | 1585 (9.0) | 2221 (8.8) | 1960 (3.3) | 10260 (6.0) |
| Numbers of IPD visits, no. (%) |  |  |  |  |  |
| 0 | 62276 (90.4) | 15831 (89.9) | 23123 (91.1) | 57596 (95.6) | 158826 (92.3) |
| 1 | 4101 (6.0) | 1132 (6.4) | 1462 (5.8) | 1709 (2.8) | 8404 (4.9) |
| 2 | 1191 (1.7) | 306 (1.7) | 400 (1.6) | 486 (0.8) | 2383 (1.4) |
| ≥3 | 1350 (2.0) | 336 (1.9) | 387 (1.5) | 450 (0.7) | 2523 (1.5) |
| Numbers of OPD visits, no. (%) |  |  |  |  |  |
| 0 | 2315 (3.4) | 643 (3.7) | 1374 (5.4) | 4498 (7.5) | 8830 (5.1) |
| 1 | 6570 (9.5) | 1751 (9.9) | 2409 (9.5) | 4544 (7.5) | 15274 (8.9) |
| 2 | 8355 (12.1) | 2105 (12.0) | 2866 (11.3) | 5929 (9.8) | 19255 (11.2) |
| ≥3 | 51678 (75.0) | 13106 (74.4) | 18723 (73.8) | 45270 (75.1) | 128777 (74.8) |
| Numbers of medications or prescriptions, no. (%) |  |  |  |  |  |
| 0 | 9944 (14.4) | 2235 (12.7) | 3878 (15.3) | 12364 (20.5) | 28421 (16.5) |
| 1 | 7759 (11.3) | 1787 (10.2) | 2627 (10.4) | 7078 (11.7) | 19251 (11.2) |
| 2 | 6998 (10.2) | 1639 (9.3) | 2404 (9.5) | 6591 (10.9) | 17632 (10.2) |
| 2+ | 44217 (64.2) | 11944 (67.8) | 16463 (64.9) | 34208 (56.8) | 106832 (62.1) |
| Dosage of vaccine, no. (%) |  |  |  |  |  |
| 0 | 53946 (78.3) | 13975 (79.4) | 20914 (82.4) | 46248 (76.8) | 135083 (78.5) |
| 1 | 2519 (3.7) | 620 (3.5) | 901 (3.6) | 2576 (4.3) | 6616 (3.8) |
| 2 | 9979 (14.5) | 2477 (14.1) | 3065 (12.1) | 9308 (15.5) | 24829 (14.4) |
| ≥3 | 2474 (3.6) | 533 (3.0) | 492 (1.9) | 2109 (3.5) | 5608 (3.3) |

Abbreviations: AAPI, Asian American/Pacific Islander; BMI, body mass index; ED, Emergency department; IPD, Inpatient department; NHB, Non-Hispanic Black; NHW, Non-Hispanic White; OPD, Outpatient department; PMCA, Pediatric Medical Complexity; SD, Standard deviation.
<sup>a</sup> Percentages may not total 100 because of rounding.

During the follow-up period, 1402 (0.8%) participants were diagnosed as PASC (U09.9), of which 751 (53.6%) were obese or severely obese; and 74317 (43.2%) participants had at least one incident occurrences of PASC symptoms and conditions, of which 38006 (51.1%) were obese or severely obese. The median of the total number of incident occurrences of PASC symptoms and conditions was 0, signifying that at least half of the participants in the cohort did not experience any incident occurrences of PASC symptoms and conditions.

Participants classified as having overweight, obesity, or severe obesity exhibited an increased risk of PASC compared to those with healthy weight, albeit not all reaching statistical significance. Specifically, in comparison to those with healthy weight, participants categorized as having obesity and severe obesity demonstrated a noteworthy 25.4% (RR, 1.254; 95% CI, 1.064-1.478) and 42.1% (RR, 1.421; 95% CI, 1.253-1.611) higher risk of PASC (U09.9), respectively. Similarly, those who had obesity and severe obesity experienced a 10.8% (RR, 1.108; 95% CI, 1.064-1.154) and 17.4% (RR, 1.174; 95% CI, 1.138-1.213) increased likelihood of encountering any manifestation of potential PASC symptoms and conditions, respectively. Furthermore, when assessing the cumulative occurrences of PASC symptoms and conditions, the association became slightly more pronounced and the RRs for those who had overweight, obesity, and severe obesity were 1.053 (95% CI, 1.000-1.109), 1.137 (95% CI, 1.088-1.188), and 1.182 (95% CI, 1.142-1.223), respectively. A significant dose-response relationship emerged between worsening BMI classification and risk of PASC (*P* < 0.05 from the linear trend test) (Table 2).

**Table 2.**
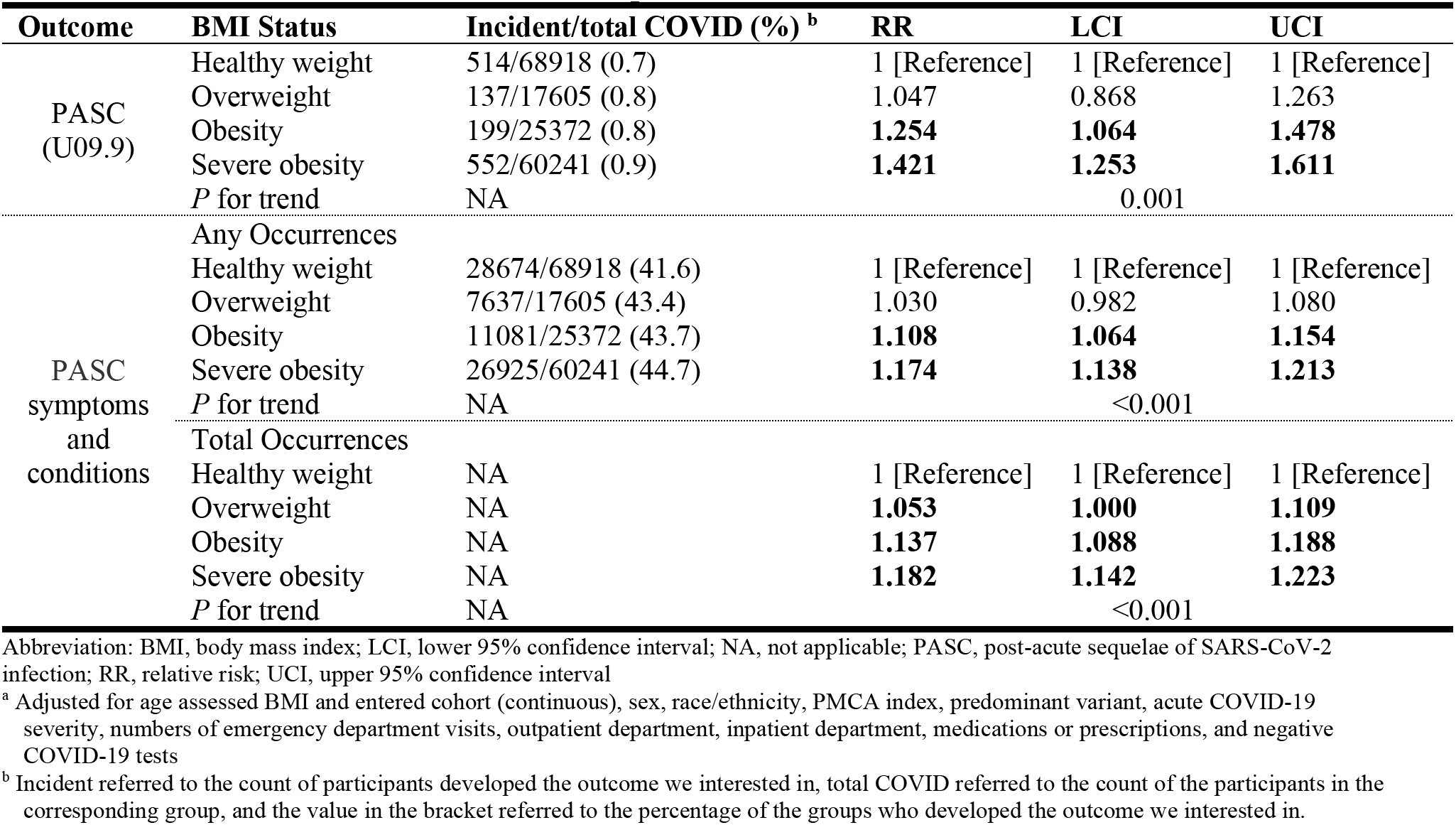
Estimated association of BMI status prior to the SARS-CoV-2 infection and risk of PASC ^a^.

Similar association was identified in the subgroup analysis by age and PMAC index, and the result did not differ significantly in the analysis for Non-Hispanic White, yet the association was not observed in the PASC (U09.9) for Non-Hispanic Black, neither was in all the PASC outcomes for Hispanic (Table 3), where the analysis for American/Pacific Islander was not present because the case of U09.9 diagnosis in certain BMI status categories was too small. The significant dose-response relationship still held in the sensitivity analysis although slightly changes were seen in the associations (eTables 4-14 in the Supplement). The result for negative control outcome analysis was insignificant, indicating that residual bias was not observed (eTable 15 in the Supplement).

**Table 3.** Estimated association of BMI status prior to the SARS-CoV-2 infection and risk of PASC in subgroups based on age, PMAC index, and race and ethnicity.

| Outcome | BMI Status | Incident/total COVID (%)<sup>a</sup> | RR | LCI | UCI |
| --- | --- | --- | --- | --- | --- |
| **Age, <18 yr (N=145975)<sup>b, c</sup>** |  |  |  |  |  |
| PASC<br>(U09.9) | Healthy weight | 408/57916 (0.7) | 1 [Reference] | 1 [Reference] | 1 [Reference] |
|  | Overweight | 110/14994 (0.7) | 1.052 | 0.854 | 1.298 |
|  | Obesity | 173/23198 (0.7) | **1.279** | **1.070** | **1.528** |
|  | Severe obesity | 424/49867 (0.9) | **1.429** | **1.238** | **1.648** |
|  | *P* for trend | NA |  | 0.008 |  |
| PASC<br>symptoms<br>and<br>conditions | Any Occurrences |  |  |  |  |
|  | Healthy weight | 23860/57916 (41.2) | 1 [Reference] | 1 [Reference] | 1 [Reference] |
|  | Overweight | 6409/14994 (42.7) | 1.020 | 0.970 | 1.074 |
|  | Obesity | 9991/23198 (43.1) | **1.099** | **1.053** | **1.146** |
|  | Severe obesity | 21923/49867 (44.0) | **1.171** | **1.131** | **1.212** |
|  | *P* for trend | NA |  | <0.001 |  |
|  | Total Occurrences |  |  |  |  |
|  | Healthy weight | NA | 1 [Reference] | 1 [Reference] | 1 [Reference] |
|  | Overweight | NA | 1.041 | 0.985 | 1.100 |
|  | Obesity | NA | **1.121** | **1.070** | **1.173** |
|  | Severe obesity | NA | **1.181** | **1.137** | **1.226** |
|  | *P* for trend | NA |  | <0.001 |  |
| **Age, ≥18 yr (N=26161)<sup>b, c</sup>** |  |  |  |  |  |
| PASC<br>(U09.9) | Healthy weight | 106/11002 (1.0) | 1 [Reference] | 1 [Reference] | 1 [Reference] |
|  | Overweight | 27/2611 (1.0) | 1.014 | 0.663 | 1.552 |
|  | Obesity | 26/2174 (1.2) | **1.142** | **0.742** | **1.758** |
|  | Severe obesity | 128/10374 (1.2) | **1.367** | **1.047** | **1.784** |
|  | *P* for trend | NA |  | 0.049 |  |
| PASC<br>symptoms<br>and<br>conditions | Any Occurrences |  |  |  |  |
|  | Healthy weight | 4814/11002 (43.8) | 1 [Reference] | 1 [Reference] | 1 [Reference] |
|  | Overweight | 1228/2611 (47.0) | 1.101 | 0.967 | 1.254 |
|  | Obesity | 1090/2174 (50.1) | **1.196** | **1.048** | **1.365** |
|  | Severe obesity | 5002/10374 (48.2) | **1.201** | **1.107** | **1.302** |
|  | *P* for trend | NA |  | <0.001 |  |
|  | Total Occurrences |  |  |  |  |
|  | Healthy weight | NA | 1 [Reference] | 1 [Reference] | 1 [Reference] |
|  | Overweight | NA | 1.140 | 0.987 | 1.317 |
|  | Obesity | NA | **1.304** | **1.124** | **1.512** |
|  | Severe obesity | NA | **1.191** | **1.090** | **1.302** |
|  | *P* for trend | NA |  | 0.002 |  |
| **PMAC index, no chronic condition (N=105294)<sup>d</sup>** |  |  |  |  |  |
| PASC<br>(U09.9) | Healthy weight | 316/41656 (0.8) | 1 [Reference] | 1 [Reference] | 1 [Reference] |
|  | Overweight | 73/10071 (0.7) | 0.965 | 0.749 | 1.243 |
|  | Obesity | 112/14427 (0.8) | **1.260** | **1.015** | **1.565** |
|  | Severe obesity | 353/39140 (0.9) | **1.456** | **1.240** | **1.708** |
|  | *P* for trend | NA |  | 0.023 |  |
| PASC<br>symptoms<br>and<br>conditions | Any Occurrences |  |  |  |  |
|  | Healthy weight | 14914/41656 (35.8) | 1 [Reference] | 1 [Reference] | 1 [Reference] |
|  | Overweight | 3718/10071 (36.9) | 0.999 | 0.944 | 1.058 |
|  | Obesity | 5423/14427 (37.6) | **1.116** | **1.064** | **1.170** |
|  | Severe obesity | 15488/39140 (39.6) | **1.184** | **1.141** | **1.228** |
|  | *P* for trend | NA |  | <0.001 |  |
|  | Total Occurrences |  |  |  |  |
|  | Healthy weight | NA | 1 [Reference] | 1 [Reference] | 1 [Reference] |
|  | Overweight | NA | 1.021 | 0.960 | 1.085 |
|  | Obesity | NA | **1.144** | **1.087** | **1.205** |
|  | Severe obesity | NA | **1.195** | **1.149** | **1.243** |
|  | *P* for trend | NA |  | <0.001 |  |
| **PMAC index, non-complex/complex chronic condition (N=66842)<sup>d</sup>** |  |  |  |  |  |
| PASC<br>(U09.9) | Healthy weight | 198/27262 (0.7) | 1 [Reference] | 1 [Reference] | 1 [Reference] |
|  | Overweight | 64/7534 (0.8) | 1.168 | 0.883 | 1.546 |
|  | Obesity | 87/10945 (0.8) | 1.255 | 0.975 | 1.615 |
|  | Severe obesity | 199/21101 (0.9) | **1.391** | **1.135** | **1.705** |
|  | *P* for trend | NA |  | 0.013 |  |
| PASC<br>symptoms<br>and<br>conditions | Any Occurrences |  |  |  |  |
|  | Healthy weight | 13760/27262 (50.5) | 1 [Reference] | 1 [Reference] | 1 [Reference] |
|  | Overweight | 3919/7534 (52.0) | **1.107** | **1.016** | **1.207** |
|  | Obesity | 5658/10945 (51.7) | **1.091** | **1.011** | **1.178** |
|  | Severe obesity | 11437/21101 (54.2) | **1.159** | **1.086** | **1.236** |
|  | *P* for trend | NA |  | <0.001 |  |
|  | Total Occurrences |  |  |  |  |
|  | Healthy weight | NA | 1 [Reference] | 1 [Reference] | 1 [Reference] |
|  | Overweight | NA | **1.132** | **1.029** | **1.246** |
|  | Obesity | NA | **1.121** | **1.031** | **1.218** |
|  | Severe obesity | NA | **1.156** | **1.077** | **1.240** |
|  | *P* for trend | NA |  | <0.001 |  |
| **Race/ethnicity, Non-Hispanic White (N=87275)<sup>e</sup>** |  |  |  |  |  |
| PASC<br>(U09.9) | Healthy weight | 371/42285 (0.9) | 1 [Reference] | 1 [Reference] | 1 [Reference] |
|  | Overweight | 83/9393 (0.9) | 0.960 | 0.758 | 1.216 |
|  | Obesity | 120/10978 (1.1) | **1.298** | **1.058** | **1.592** |
|  | Severe obesity | 333/24619 (1.4) | **1.548** | **1.332** | **1.799** |
|  | *P* for trend | NA |  | <0.001 |  |
| PASC<br>symptoms<br>and<br>conditions | Any Occurrences |  |  |  |  |
|  | Healthy weight | 17827/42285 (42.2) | 1 [Reference] | 1 [Reference] | 1 [Reference] |
|  | Overweight | 4208/9393 (44.8) | 1.022 | 0.958 | 1.091 |
|  | Obesity | 5090/10978 (46.4) | **1.074** | **1.011** | **1.142** |
|  | Severe obesity | 12363/24619 (50.2) | **1.224** | **1.171** | **1.280** |
|  | *P* for trend | NA |  | <0.001 |  |
|  | Total Occurrences |  |  |  |  |
|  | Healthy weight | NA | 1 [Reference] | 1 [Reference] | 1 [Reference] |
|  | Overweight | NA | 1.039 | 0.968 | 1.115 |
|  | Obesity | NA | **1.105** | **1.034** | **1.182** |
|  | Severe obesity | NA | **1.229** | **1.172** | **1.290** |
|  | *P* for trend | NA |  | <0.001 |  |
| **Race/ethnicity, Hispanic (N=42982)<sup>e</sup>** |  |  |  |  |  |
| PASC<br>(U09.9) | Healthy weight | 70/10657 (0.7) | 1 [Reference] | 1 [Reference] | 1 [Reference] |
|  | Overweight | 27/3588 (0.8) | 1.108 | 0.712 | 1.725 |
|  | Obesity | 48/7335 (0.7) | 1.138 | 0.786 | 1.648 |
|  | Severe obesity | 129/21402 (0.6) | 1.020 | 0.757 | 1.374 |
|  | *P* for trend | NA |  | 0.446 |  |
| PASC<br>symptoms<br>and<br>conditions | Any Occurrences |  |  |  |  |
|  | Healthy weight | 4758/10657 (44.6) | 1 [Reference] | 1 [Reference] | 1 [Reference] |
|  | Overweight | 1634/3588 (45.5) | 1.016 | 0.918 | 1.125 |
|  | Obesity | 3184/7335 (43.4) | 1.045 | 0.959 | 1.140 |
|  | Severe obesity | 8591/21402 (40.1) | **1.154** | **1.073** | **1.241** |
|  | *P* for trend | NA |  | 0.684 |  |
|  | Total Occurrences |  |  |  |  |
|  | Healthy weight | NA | 1 [Reference] | 1 [Reference] | 1 [Reference] |
|  | Overweight | NA | 1.021 | 0.914 | 1.139 |
|  | Obesity | NA | 1.076 | 0.980 | 1.182 |
|  | Severe obesity | NA | **1.167** | **1.078** | **1.263** |
|  | *P* for trend | NA |  | 0.638 |  |
| **Race/ethnicity, Non-Hispanic Black (N=33065) <sup>e</sup>** |  |  |  |  |  |
| PASC<br>(U09.9) | Healthy weight | 53/11980 (0.4) | 1 [Reference] | 1 [Reference] | 1 [Reference] |
|  | Overweight | 21/3854 (0.5) | 1.210 | 0.729 | 2.008 |
|  | Obesity | 23/6004 (0.4) | 0.916 | 0.563 | 1.492 |
|  | Severe obesity | 66/11227 (0.6) | 1.326 | 0.908 | 1.937 |
|  | *P* for trend | NA |  | 0.204 |  |
| PASC<br>symptoms<br>and<br>conditions | Any Occurrences |  |  |  |  |
|  | Healthy weight | 4665/11980 (38.9) | 1 [Reference] | 1 [Reference] | 1 [Reference] |
|  | Overweight | 1514/3854 (39.3) | 1.016 | 0.918 | 1.125 |
|  | Obesity | 2370/6004 (39.5) | 1.045 | 0.959 | 1.140 |
|  | Severe obesity | 4827/11227 (43.0) | **1.154** | **1.073** | **1.241** |
|  | *P* for trend | NA |  | <0.001 |  |
|  | Total Occurrences |  |  |  |  |
|  | Healthy weight | NA | 1 [Reference] | 1 [Reference] | 1 [Reference] |
|  | Overweight | NA | 1.021 | 0.914 | 1.139 |
|  | Obesity | NA | 1.076 | 0.980 | 1.182 |
|  | Severe obesity | NA | **1.167** | **1.078** | **1.263** |
|  | *P* for trend | NA |  | 0.002 |  |

Abbreviation: BMI, body mass index; LCI, lower 95% confidence interval; NA, not applicable; PASC, post-acute sequelae of SARS-CoV-2 infection; RR, relative risk; UCI, upper 95% confidence interval
<sup>a</sup> Incident referred to the count of participants developed the outcome we interested in, total COVID referred to the count of the participants in the corresponding group, and the value in the bracket referred to the percentage of the groups who developed the outcome we interested in.
<sup>b</sup> Adjusted for age assessed BMI and entered cohort (continuous), sex, race/ethnicity, PMCA index, predominant variant, acute COVID-19 severity, numbers of emergency department visits, outpatient department, inpatient department, medications or prescriptions, and negative COVID-19 tests
<sup>c</sup> Stratified by the cohort entry age.
<sup>d</sup> Adjusted for age assessed BMI and entered cohort (continuous), sex, predominant variant, acute COVID-19 severity, numbers of emergency department visits, outpatient department, inpatient department, medications or prescriptions, and negative COVID-19 tests
<sup>e</sup> Adjusted for age assessed BMI and entered cohort (continuous), sex, PMCA index, predominant variant, acute COVID-19 severity, numbers of emergency department visits, outpatient department, inpatient department, medications or prescriptions, and negative COVID-19 tests

## Discussion

To our knowledge, this retrospective cohort study is the first and the largest to explore the relationship between BMI status and PASC among the pediatric population. Within the follow-up period, an adverse dose-response relationship between pre-infection BMI status and the susceptibility to PASC was consistently revealed, after adjusting for sociodemographic and clinical variables. This association pattern between BMI status and PASC risk was consistent across pediatric and adult populations^19^, albeit with varying magnitudes of association. This association persisted even after rigorous adjustments for critical factors through secondary analyses, except for the subgroup analysis by racial and ethnic minorities, suggesting that race/ethnicity may be an effect modifier. Importantly, the incorporation of negative control outcome yielded no sizable residual bias, confirming the robustness and reliability of the findings.

Our study had several strengths. A pivotal one lies in its extensive and representative sample. Covering 172136 pediatric populations with COVID-19 infection from twenty-six US children’s hospitals, our study is distinguished by its scale, enabling us ample statistical power to rigorously evaluate the BMI-PASC relationship accounting for key sociodemographic and clinical risk factors. A noteworthy distinction from the previous research is our ICD-coded EHR phenotypes rather than self-reported PASC symptoms as the latter may be influenced by individual perceptions, interpretations, recall biases, lack of standardization, participant compliance with reporting protocols, etc. The inclusion of multiple PASC outcomes enhances the clarity in understanding the burden and risk of PASC, and mitigates the potential for misclassification. This is particularly relevant given the recognized limitations associated with only using U09.9^36^ and acknowledging the likely divergence in clinical features of PASC between pediatric and adult population.^23^ Moreover, a considerable portion of the existing studies primarily focused solely on hospitalized cohorts, our cohort is characterized by its inclusivity, incorporating both non-hospitalized and hospitalized participants.

Although our study revealed a significantly higher obesity rate (35.7% vs. 49.7% before and after excluding those who were underweight and missed BMI status) compared to the national average (19.7%),^15^ the rate among participants from primary care sites was comparable (17.52% vs. 22.36% before and after excluding those who were underweight and missed BMI status). This disparity may be attributed to better overall health among participants from primary care sites, as well as the heightened susceptibility to various health conditions among overweight or obese individuals.^37^ Higher BMI has also been associated with increased risks of hospitalization^38^ and severe illness^39^ among pediatric COVID-19 patients. Alternatively, it is possible that pediatric BMI levels, particularly in the overweight^29^ and obesity^40^ categories, increased during the pandemic, which could explain the increased RRs observed for individual in these categories in the sensitivity analysis, which excluded participants whose BMI assessment date was beyond 6 months before cohort entry. The BMI-PASC association remained consistent across the outcome measures, although stronger associations were observed for U09.9. The differences observed could stem from the specificity of PASC symptoms and conditions compared to U09.9. Recognizing the limitations associated with the use of U09.9^36^ and PASC symptoms and conditions,^23^ we incorporated both measures to enhance the comprehensiveness and precision in our analysis.

The associations observed in our study may be explained by several plausible biological mechanisms, although they may not be pediatric-specific. Firstly, obesity’s association with chronic inflammation ^41,42^ and susceptibility to COVID-19 may involve macrophages pyroptosis,^43^ leading to prolonged systemic inflammation implicated in the genesis of PASC.^44,45^ Secondly, obesity is also associated with altered microbiota,^46^ and the impact of COVID-19 on the latter may influence PASC risk.^47^ Thirdly, obesity is recognized for its propensity to dysregulate adaptive autoimmunity,^48,49^ a phenomenon documented in PASC patients.^50,51^ This dysregulation in adaptive immune responses may be implicated in the protracted nature of observed symptoms. Furthermore, clotting and endothelial abnormalities associated with obesity could explain observed pathophysiological changes in PASC patients.^50,52–54^

Our study has several limitations. Firstly, the high prevalence of obesity among participants may indicate sample skewness, which can be explained by several factors, including the missing BMI status data within the baseline period, as discussed above, thus necessitating cautious generalization to the entire US pediatric population. Additionally, misclassification bias challenges pediatric PASC diagnosis due to the lack of standardized reference. To mitigate this, we used ICD-10-CM codes and PASC symptoms and conditions, though the latter may not capture all aspects. Moreover, some PASC symptoms and conditions we examined may overlap with those in pediatric populations with obesity, potentially inflating the association. However, we assessed the incidence of these symptoms and conditions do not present during the baseline period, which may alleviate this issue. To address the probable underestimation of the PASC burden, we included participants with at least one visit two years before cohort entry and one during the follow-up period. Furthermore, the absence of information on modifiable risk factors, such as diet, physical activity, or sleep in our dataset is notable. Previous study has linked these factors to a substantially reduced PASC risk.^19^ Therefore, further investigations including these modifiable factors involving the pediatric population are warranted.

## Conclusion

Our study reveals a significant association between higher pre-SARS-CoV-2 infection BMI and increased PASC risk in pediatric patients, which contributes to the understanding and management of PASC in this context. Given the potential for associated PASC conditions to become lifelong chronic conditions, understanding pediatric PASC is important. Clinically, this association emphasizes the need for vigilant monitoring and tailored management for this demographic with elevated BMI who have had COVID-19. Pediatricians play a vital role in identifying at-risk individuals and implementing personalized treatment plans to mitigate long-term consequences. From a public health standpoint, recognizing the connection between elevated BMI and PASC highlights the importance of preventive interventions. Public health initiatives can raise awareness about the heightened PASC risk among obese pediatrics, promoting healthy lifestyle behaviors to reduce severe COVID-19 outcomes. Collaborative efforts are essential for implementing programs that promote healthy weight management and foster supportive environments. Addressing obesity as a modifiable risk factor for pediatric PASC can alleviate the burden of PASC and further the broader goals of pediatric health post-pandemic. Further research should explore specific PASC symptoms and conditions across various organ systems.

## Data Availability

All data produced in the present work are contained in the manuscript.

## Disclosures

### Disclaimer

This content is solely the responsibility of the authors and does not necessarily represent the official views of the RECOVER Initiative, the NIH, or other funders.

## Acknowledgements

This study is part of the NIH Researching COVID-19 to Enhance Recovery (RECOVER) Initiative, which seeks to understand, treat, and prevent the post-acute sequelae of SARS-CoV-2 infection (PASC). For more information on RECOVER, visit https://recovercovid.org/.

We would like to thank the National Community Engagement Group (NCEG), all patients, caregivers, and community Representatives, and all the participants enrolled in the RECOVER Initiative. We want to thank to Max Hornig, Teresa Akintonwa, Etienne Carignan for their helpful suggestions on our manuscript.

