## Supplement for "Overweight and Obesity as Predictors of Post-acute Sequelae of SARS-Cov-2 Infection: Findings from the RECOVER Initiative": Supplement_Overweight and Obesity as Predictors of Post-acute Sequelae of SARS-Cov-2 Infection.pdf

**eFigure 1.** The participants selection process

**eTable 1.** The twenty-six sites that contributed data in the analysis

**eTable 2.** Details for the medical conditions in the exclusion criteria

**eTable 3.** Characteristics according to missingness of BMI status prior to COVID-19 infection

**eTable 4.** Estimated association of BMI status prior to the SARS-CoV-2 infection and risk of PASC by considering the assessment time of BMI status [a] (N=105 326)

**eTable 5.** Estimated association of BMI status prior to the SARS-CoV-2 infection and risk of PASC by considering the release time of U09.9 [a] (N=101 143)

**eTable 6.** Estimated association of BMI status prior to the SARS-CoV-2 infection and risk of PASC after excluding participants confirmed by serology test after November 2022 (N=171 978)

**eTable 7.** Estimated association of BMI status prior to the SARS-CoV-2 infection and risk of after excluding severe or moderate participants (N=163 094)

**eTable 8.** Estimated association of BMI status prior to the SARS-CoV-2 infection and risk of PASC adjusting for doses of COVID-19 vaccine before infection (N=172 316)

**eTable 9.** Estimated association of BMI status prior to the SARS-CoV-2 infection and risk of PASC adjusting for interval since last COVID-19 vaccination date (N=172 316)

**eTable 10.** Estimated association of BMI status prior to the SARS-CoV-2 infection and risk of PASC adjusting for doses of COVID-19 vaccine before infection and interval since last COVID-19 vaccination date (N=172 316)

**eTable 11.** Estimated association of BMI status prior to the SARS-CoV-2 infection and risk of PASC adjusting type of insurance[1] (N=140 036)

**eTable 12.** Estimated association of BMI status prior to the SARS-CoV-2 infection and risk of PASC after excluding participants with diabetes (N=169 495)

**eTable 13.** Estimated association of BMI status prior to the SARS-CoV-2 infection and risk of after excluding obese participants taking weight loss drugs [a] (N=169 255)

**eTable 14.** Estimated association of BMI status prior to the SARS-CoV-2 infection and risk of PASC based on primary care sites (N=42 470)

**eTable 15.** Estimated association of BMI status prior to the SARS-CoV-2 infection and risk of foreign body in ear as a negative control outcome (N=172 316)

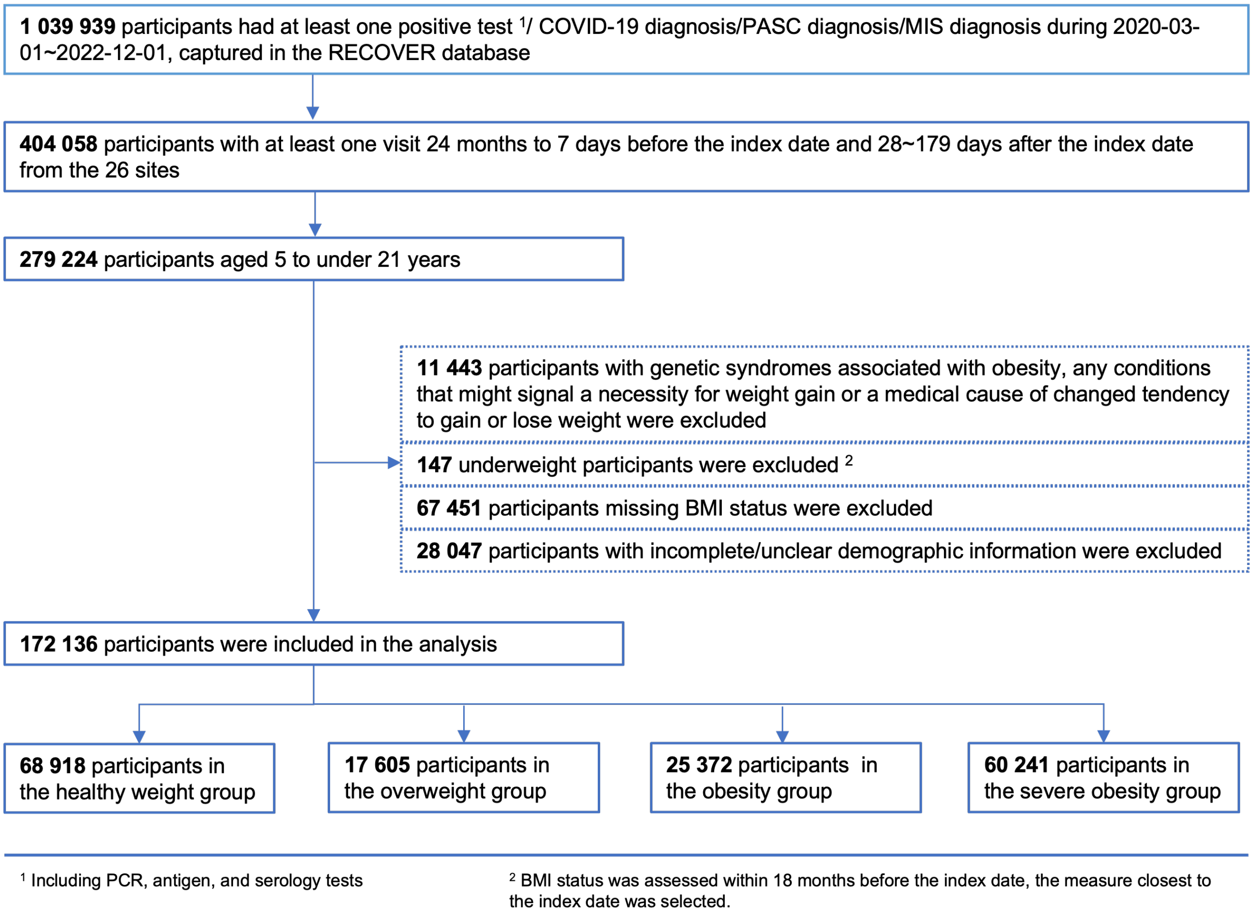


**eFigure 1. The participants selection process**

**eTable 1. The twenty-six sites that contributed data in the analysis**

| ID | Site Name |
|---|---|
| 1 | Albert Einstein College of Medicine |
| 2 | Ann & Robert H. Lurie Children’s Hospital of Chicago |
| 3 | Cincinnati Children’s Hospital Medical Center |
| 4 | Children’s Hospital of Philadelphia |
| 5 | Children’s National Hospital |
| 6 | Colorado Children’s Hospital |
| 7 | Duke University Health System |
| 8 | Medical College of Wisconsin |
| 9 | Medical University of South Carolina |
| 10 | Nationwide Children’s Hospital |
| 11 | New York University Langone Health |
| 12 | Northwestern University |
| 13 | OCHIN, Inc |
| 14 | Seattle Children’s Hospital |
| 15 | Stanford University |
| 16 | The Nemours Foundation |
| 17 | The Ohio State University |
| 18 | The University of Iowa |
| 19 | University of California San Francisco |
| 20 | University of Michigan |
| 21 | University of Missouri |
| 22 | University of Nebraska Medical Center |
| 23 | UPMC-Children's Hospital of Pittsburgh |
| 24 | Vanderbilt University Medical Center |
| 25 | Wake Forest University Health Sciences |
| 26 | Weill Cornell Medicine |

### eTable 2. Details for the medical conditions in the exclusion criteria

| Condition | Condition Name |
|---|---|
| Genetic syndromes associated with obesity | MC4R deficiency, Leptin deficiency, Leptin receptor deficiency, POMC deficiency, Proprotein subtilisin or kexin type 1 deficiency, SRC1 deficiency, Prader-Willi syndrome, Alstrom syndrome, Bardet-Biedl syndrome, Smith-Magenis syndrome, SH2B1 deficiency, Sim1 deficiency, 16p11.2 microdeletion syndrome, Brain derived neurotrophic factor deficiency, Albright’s hereditary osteodystrophy, Cohen syndrome, Beckwith-Wiedemann syndrome |
| Conditions signaling a need for weight gain or a medical cause of altered weight tendencies | Bariatric surgery, BMI less than $5^{th}$ percentile for age and sex, cancer, Crohn's disease, Cushing syndrome, cystic fibrosis, eating disorder, growth hormone deficiency, HIV/AIDS, panhypopituitarism, pregnancy, sickle cell disease, ulcerative colitis |

**eTable 3. Characteristics according to missingness of BMI status prior to COVID-19 infection [a]**

| | Missing BMI status | | |
|---|---|---|---|
| | Yes (N=67 451) | No (N=172 136) | *P* value [b] |
| Mean age (SD), yr [c] | 10.07 (6.41) | 13.06 (4.37) | <0.001 |
| Sex, no. (%) | | | |
| Male | 33 772 (50.07) | 81 949 (47.61) | <0.001 |
| Female | 33 679 (49.93) | 90 187 (52.39) | <0.001 |
| Race, no. (%) | | | |
| NHW | 35 156 (52.12) | 87 275 (50.70) | <0.001 |
| AAPI | 3 344 (4.96) | 8 814 (5.12) | 0.105 |
| NHB | 13 475 (19.98) | 33 065 (19.21) | <0.001 |
| Hispanic | 15 476 (22.94) | 42 982 (24.97) | <0.001 |
| Predominant variant, no. (%) | | | |
| Pre-Alpha | 17 496 (25.94) | 45 440 (26.40) | 0.022 |
| Alpha | 3 795 (5.63) | 8 555 (4.97) | <0.001 |
| Delta | 17 527 (25.98) | 42 490 (24.68) | <0.001 |
| Omicron | 28 633 (42.45) | 75 651 (43.95) | <0.001 |
| PMCA, no. (%) | | | |
| None | 58 732 (87.07) | 105 294 (61.17) | <0.001 |
| Noncomplex | 6 304 (9.35) | 40 135 (23.32) | <0.001 |
| Complex | 2 415 (3.58) | 26 707 (15.52) | <0.001 |
| Severity, no. (%) | | | |
| Asymptomatic | 40 799 (60.49) | 102 366 (59.47) | <0.001 |
| Mild | 21 787 (32.30) | 60 728 (35.28) | <0.001 |
| Moderate | 3 305 (4.90) | 5 704 (3.31) | <0.001 |
| Severe | 1 560 (2.31) | 3 338 (1.94) | <0.001 |
| Numbers of negative COVID-19 tests, no. (%) | | | |
| 0 | 43 079 (63.87) | 104 312 (60.60) | <0.001 |
| 1 | 14 719 (21.82) | 36 964 (21.47) | 0.063 |
| ≥2 | 9 653 (14.31) | 30 860 (17.93) | <0.001 |
| Numbers of ED visits, no. (%) | | | |
| 0 | 46 010 (68.21) | 129 862 (75.44) | <0.001 |
| 1 | 12 164 (18.03) | 22 896 (13.30) | <0.001 |
| 2 | 4 670 (6.92) | 9 188 (5.30) | <0.001 |
| ≥3 | 4 670 (6.83) | 10 260 (5.96) | <0.001 |
| Numbers of IPD visits, no. (%) | | | |
| 0 | 65 195 (96.66) | 158 826 (92.27) | <0.001 |
| 1 | 1 912 (2.83) | 8 404 (4.88) | <0.001 |
| 2 | 214 (0.32) | 2 383 (1.38) | <0.001 |
| ≥3 | 130 (0.19) | 2 523 (1.47) | <0.001 |
| Numbers of OPD visits, no. (%) | | | |
| 0 | 28 339 (42.01) | 8 830 (5.13) | <0.001 |
| 1 | 14 479 (21.47) | 15 274 (8.87) | <0.001 |
| 2 | 7 976 (11.82) | 19 255 (11.19) | <0.001 |
| ≥3 | 16 657 (24.69) | 128 777 (74.81) | <0.001 |
| Numbers of medications or prescriptions, no. (%) | | | |
| 0 | 31 523 (46.73) | 28 421 (16.51) | <0.001 |
| 1 | 9 749 (14.45) | 19 251 (11.18) | <0.001 |
| 2 | 6 827 (10.12) | 17 632 (10.24) | 0.381 |
| ≥3 | 19 352 (28.69 | 106 832 (62.06) | <0.001 |
| PASC (U09.9) , no. (%) | 532 (0.79) | 1 402 (0.81) | 0.543 |
| Any occurrence of PASC symptoms, no. (%) | 24 232 (35.93) | 74 317 (43.17) | <0.001 |
| Median total occurrence of PASC symptoms (IRQ) | 0 (0-14) | 0 (0-15) | <0.001 |

Abbreviations: AAPI, Asian American/Pacific Islander; BMI, body mass index; ED, Emergency department; IPD, Inpatient department; IQR, interquartile range; NHB, Non-Hispanic Black; NHW, Non-Hispanic White; OPD, Outpatient department; PASC, post-acute sequelae of SARS-CoV-2 infection; PMCA, Pediatric Medical Complexity; SD, Standard deviation.

[a] Percentages may not total 100 because of rounding.

[b] Student’s t test for age and total occurrence of PASC symptoms, contingency chi-square test for the other categorical variables.

[c] Referred to the cohort entry age.

**eTable 4. Estimated association of BMI status prior to the SARS-CoV-2 infection and risk of PASC by considering the assessment time of BMI status [ab] (N=105 326)**

| Outcome | BMI Status | Incident/total COVID (%) [c] | RR | LCI | UCI |
|---|---|---|---|---|---|
| PASC (U09.9) | Healthy weight | 338/42 404 (0.8) | 1 [Ref] | 1 [Ref] | 1 [Ref] |
| | Overweight | 100/11 049 (0.9) | 1.153 | 0.924 | 1.439 |
| | Obesity | 137/15 879 (0.9) | **1.305** | **1.069** | **1.592** |
| | Severe obesity | 352/35 994 (1.0) | **1.403** | **1.201** | **1.638** |
| | *P* for trend | NA | | 0.001 | |
| PASC symptoms and conditions | **Any Occurrences** | | | | |
| | Healthy weight | 19 292/42 404 (45.5) | 1 [Ref] | 1 [Ref] | 1 [Ref] |
| | Overweight | 5 270/11 049 (47.7) | 1.052 | 0.987 | 1.121 |
| | Obesity | 7 520/15 879 (47.4) | **1.109** | **1.050** | **1.172** |
| | Severe obesity | 17 473/35 994 (48.5) | **1.171** | **1.121** | **1.223** |
| | *P* for trend | NA | | <0.001 | |
| | **Total Occurrences** | | | | |
| | Healthy weight | NA | 1 [Ref] | 1 [Ref] | 1 [Ref] |
| | Overweight | NA | **1.099** | **1.025** | **1.179** |
| | Obesity | NA | **1.146** | **1.080** | **1.216** |
| | Severe obesity | NA | **1.186** | **1.132** | **1.244** |
| | *P* for trend | NA | | <0.001 | |

Abbreviation: BMI, body mass index; LCI, lower 95% confidence interval; NA, not applicable; PASC, post-acute sequelae of SARS-CoV-2 infection; RR, relative risk; UCI, upper 95% confidence interval

[a] The BMI status was assessed within 6 months before cohort entry rather than 18 months before cohort entry

[b] Adjusted for age assessed BMI and entered cohort (continuous), sex, race/ethnicity, PMCA index, predominant variant, acute COVID-19 severity, numbers of emergency department visits, outpatient department visits, inpatient department visits, medications or prescriptions, and negative COVID-19 tests

[c] Incident referred to the count of participants developed the outcome we interested in, total COVID referred to the count of the participants in the corresponding group, and the value in the bracket referred to the percentage of the groups who developed the outcome we interested in.

**eTable 5. Estimated association of BMI status prior to the SARS-CoV-2 infection and risk of PASC by considering the release time of U09.9 [ab] (N=101 143)**

| Outcome | BMI Status | Incident/total COVID (%) [c] | RR | LCI | UCI |
|---|---|---|---|---|---|
| PASC (U09.9) | Healthy weight | 427/40 490 (1.1) | 1 [Ref] | 1 [Ref] | 1 [Ref] |
| | Overweight | 112/10 205 (1.1) | 1.047 | 0.852 | 1.288 |
| | Obesity | 158/15 335 (1.0) | **1.218** | **1.014** | **1.462** |
| | Severe obesity | 451/35 113 (1.3) | **1.404** | **1.221** | **1.613** |
| | *P* for trend | NA | | 0.006 | |
| PASC symptoms and conditions | **Any Occurrences** | | | | |
| | Healthy weight | 17 518/40 490 (43.3) | 1 [Ref] | 1 [Ref] | 1 [Ref] |
| | Overweight | 4 642/10 205 (45.5) | 1.026 | 0.964 | 1.091 |
| | Obesity | 7 086/15 335 (46.2) | **1.147** | **1.091** | **1.207** |
| | Severe obesity | 16 659/35 113 (47.4) | **1.203** | **1.156** | **1.253** |
| | *P* for trend | NA | | <0.001 | |
| | **Total Occurrences** | | | | |
| | Healthy weight | NA | 1 [Ref] | 1 [Ref] | 1 [Ref] |
| | Overweight | NA | 1.051 | 0.982 | 1.124 |
| | Obesity | NA | **1.178** | **1.115** | **1.244** |
| | Severe obesity | NA | **1.214** | **1.162** | **1.268** |
| | *P* for trend | NA | | <0.001 | |

Abbreviation: BMI, body mass index; LCI, lower 95% confidence interval; NA, not applicable; PASC, post-acute sequelae of SARS-CoV-2 infection; RR, relative risk; UCI, upper 95% confidence interval

[a] The cohort entry date was restricted to after October 1, 2021 when the U09.9 was released.

[b] Adjusted for age assessed BMI and entered cohort (continuous), sex, race/ethnicity, PMCA index, predominant variant, acute COVID-19 severity, numbers of emergency department visits, outpatient department visits, inpatient department visits, medications or prescriptions, and negative COVID-19 tests

[c] Incident referred to the count of participants developed the outcome we interested in, total COVID referred to the count of the participants in the corresponding group, and the value in the bracket referred to the percentage of the groups who developed the outcome we interested in.

**eTable 6. Estimated association of BMI status prior to the SARS-CoV-2 infection and risk of PASC after excluding participants confirmed by serology test after November 2022 (N=171 978) [a]**

| Outcome | BMI Status | Incident/total COVID (%) [b] | RR | LCI | UCI |
|---|---|---|---|---|---|
| PASC (U09.9) | Healthy weight | 506/68 827 (0.7) | 1 [Ref] | 1 [Ref] | 1 [Ref] |
| | Overweight | 137/17 585 (0.8) | 1.064 | 0.882 | 1.284 |
| | Obesity | 196/25 333 (0.8) | **1.259** | **1.067** | **1.485** |
| | Severe obesity | 551/60 233 (0.9) | **1.437** | **1.267** | **1.630** |
| | *P* for trend | NA | | 0.001 | |
| PASC symptoms and conditions | **Any Occurrences** | | | | |
| | Healthy weight | 28 600/68 827 (41.6) | 1 [Ref] | 1 [Ref] | 1 [Ref] |
| | Overweight | 7 620/17 585 (43.3) | 1.028 | 0.981 | 1.079 |
| | Obesity | 11 050/25 333 (43.6) | **1.108** | **1.064** | **1.154** |
| | Severe obesity | 26 919/60 233 (44.7) | **1.177** | **1.140** | **1.215** |
| | *P* for trend | NA | | <0.001 | |
| | **Total Occurrences** | | | | |
| | Healthy weight | NA | 1 [Ref] | 1 [Ref] | 1 [Ref] |
| | Overweight | NA | 1.049 | 0.996 | 1.104 |
| | Obesity | NA | **1.137** | **1.088** | **1.188** |
| | Severe obesity | NA | **1.186** | **1.146** | **1.228** |
| | *P* for trend | NA | | <0.001 | |

Abbreviation: BMI, body mass index; LCI, lower 95% confidence interval; NA, not applicable; PASC, post-acute sequelae of SARS-CoV-2 infection; RR, relative risk; UCI, upper 95% confidence interval

[a] Adjusted for age assessed BMI and entered cohort (continuous), sex, race/ethnicity, PMCA index, predominant variant, acute COVID-19 severity, numbers of emergency department visits, outpatient department visits, inpatient department visits, medications or prescriptions, and negative COVID-19 tests

[b] Incident referred to the count of participants developed the outcome we interested in, total COVID referred to the count of the participants in the corresponding group, and the value in the bracket referred to the percentage of the groups who developed the outcome we interested in.

**eTable 7. Estimated association of BMI status prior to the SARS-CoV-2 infection and risk of PASC after excluding severe or moderate participants (N=163 094) [a]**

| Outcome | BMI Status | Incident/total COVID (%) [b] | RR | LCI | UCI |
|---|---|---|---|---|---|
| PASC (U09.9) | Healthy weight | 476/64 674 (0.7) | 1 [Ref] | 1 [Ref] | 1 [Ref] |
| | Overweight | 131/16 482 (0.8) | 1.095 | 0.903 | 1.326 |
| | Obesity | 186/23 837 (0.8) | **1.243** | **1.049** | **1.473** |
| | Severe obesity | 526/58 101 (0.9) | **1.282** | **1.128** | **1.456** |
| | *P* for trend | NA | | 0.001 | |
| PASC symptoms and conditions | **Any Occurrences** | | | | |
| | Healthy weight | 26 269/64 674 (40.6) | 1 [Ref] | 1 [Ref] | 1 [Ref] |
| | Overweight | 6 991/16 482 (42.4) | 1.022 | 0.973 | 1.074 |
| | Obesity | 10 246/23 837 (43.0) | **1.116** | **1.070** | **1.163** |
| | Severe obesity | 25 631/58 101 (44.1) | **1.219** | **1.180** | **1.260** |
| | *P* for trend | NA | | <0.001 | |
| | **Total Occurrences** | | | | |
| | Healthy weight | NA | 1 [Ref] | 1 [Ref] | 1 [Ref] |
| | Overweight | NA | 1.039 | 0.986 | 1.096 |
| | Obesity | NA | **1.148** | **1.097** | **1.201** |
| | Severe obesity | NA | **1.232** | **1.190** | **1.276** |
| | *P* for trend | NA | | <0.001 | |

Abbreviation: BMI, body mass index; LCI, lower 95% confidence interval; NA, not applicable; PASC, post-acute sequelae of SARS-CoV-2 infection; RR, relative risk; UCI, upper 95% confidence interval

[a] Adjusted for age assessed BMI and entered cohort (continuous), sex, race/ethnicity, PMCA index, predominant variant, numbers of emergency department visits, outpatient department visits, inpatient department visits, medications or prescriptions, and negative COVID-19 tests

[b] Incident referred to the count of participants developed the outcome we interested in, total COVID referred to the count of the participants in the corresponding group, and the value in the bracket referred to the percentage of the groups who developed the outcome we interested in.

**eTable 8. Estimated association of BMI status prior to the SARS-CoV-2 infection and risk of PASC adjusting for doses of COVID-19 vaccine before infection (N=172 316) [a]**

| Outcome | BMI Status | Incident/total COVID (%) [b] | RR | LCI | UCI |
|---|---|---|---|---|---|
| PASC (U09.9) | Healthy weight | 514/68 918 (0.7) | 1 [Ref] | 1 [Ref] | 1 [Ref] |
| | Overweight | 137/17 605 (0.8) | 1.046 | 0.868 | 1.262 |
| | Obesity | 199/25 372 (0.8) | **1.251** | **1.062** | **1.475** |
| | Severe obesity | 552/60 241 (0.9) | **1.422** | **1.250** | **1.611** |
| | *P* for trend | NA | | 0.001 | |
| PASC symptoms and conditions | **Any Occurrences** | | | | |
| | Healthy weight | 28 674/68 918 (41.6) | 1 [Ref] | 1 [Ref] | 1 [Ref] |
| | Overweight | 7 637/17 605 (43.4) | 1.025 | 0.973 | 1.080 |
| | Obesity | 11 081/25 372 (43.7) | **1.072** | **1.025** | **1.121** |
| | Severe obesity | 26 925/60 241 (44.7) | **1.125** | **1.086** | **1.165** |
| | *P* for trend | NA | | <0.001 | |
| | **Total Occurrences** | | | | |
| | Healthy weight | NA | 1 [Ref] | 1 [Ref] | 1 [Ref] |
| | Overweight | NA | 1.054 | 0.996 | 1.115 |
| | Obesity | NA | **1.099** | **1.047** | **1.153** |
| | Severe obesity | NA | **1.133** | **1.090** | **1.177** |
| | *P* for trend | NA | | <0.001 | |

Abbreviation: BMI, body mass index; LCI, lower 95% confidence interval; NA, not applicable; PASC, post-acute sequelae of SARS-CoV-2 infection; RR, relative risk; UCI, upper 95% confidence interval

[a] Adjusted for age assessed BMI and entered cohort (continuous), sex, race/ethnicity, PMCA index, predominant variant, acute COVID-19 severity, numbers of emergency department visits, outpatient department visits, inpatient department visits, medications or prescriptions, negative COVID-19 tests, and doses of COVID-19 vaccine before infection.

[b] Incident referred to the count of participants developed the outcome we interested in, total COVID referred to the count of the participants in the corresponding group, and the value in the bracket referred to the percentage of the groups who developed the outcome we interested in.

**eTable 9. Estimated association of BMI status prior to the SARS-CoV-2 infection and risk of PASC adjusting for interval since last COVID-19 vaccination date (N=172 316)[a]**

| Outcome | BMI Status | Incident/total COVID (%)[b] | RR | LCI | UCI |
|---|---|---|---|---|---|
| PASC (U09.9) | Healthy weight | 514/68 918 (0.7) | 1 [Ref] | 1 [Ref] | 1 [Ref] |
| | Overweight | 137/17 605 (0.8) | 1.046 | 0.867 | 1.261 |
| | Obesity | 199/25 372 (0.8) | **1.249** | **1.060** | **1.472** |
| | Severe obesity | 552/60 241 (0.9) | **1.420** | **1.253** | **1.610** |
| | *P* for trend | NA | | 0.001 | |
| PASC symptoms and conditions | **Any Occurrences** | | | | |
| | Healthy weight | 28 674/68 918 (41.6) | 1 [Ref] | 1 [Ref] | 1 [Ref] |
| | Overweight | 7 637/17 605 (43.4) | 1.029 | 0.981 | 1.079 |
| | Obesity | 11 081/25 372 (43.7) | **1.106** | **1.062** | **1.152** |
| | Severe obesity | 26 925/60 241 (44.7) | **1.175** | **1.138** | **1.213** |
| | *P* for trend | NA | | <0.001 | |
| | **Total Occurrences** | | | | |
| | Healthy weight | NA | 1 [Ref] | 1 [Ref] | 1 [Ref] |
| | Overweight | NA | 1.052 | 0.999 | 1.108 |
| | Obesity | NA | **1.135** | **1.086** | **1.186** |
| | Severe obesity | NA | **1.182** | **1.142** | **1.223** |
| | *P* for trend | NA | | <0.001 | |

Abbreviation: BMI, body mass index; LCI, lower 95% confidence interval; NA, not applicable; PASC, post-acute sequelae of SARS-CoV-2 infection; RR, relative risk; UCI, upper 95% confidence interval

[a] Adjusted for age assessed BMI and entered cohort (continuous), sex, race/ethnicity, PMCA index, predominant variant, acute COVID-19 severity, numbers of emergency department visits, outpatient department visits, inpatient department visits, medications or prescriptions, negative COVID-19 tests, interval since last COVID-19 vaccination date.

[b] Incident referred to the count of participants developed the outcome we interested in, total COVID referred to the count of the participants in the corresponding group, and the value in the bracket referred to the percentage of the groups who developed the outcome we interested in.

**eTable 10. Estimated association of BMI status prior to the SARS-CoV-2 infection and risk of PASC adjusting for doses of COVID-19 vaccine before infection and interval since last COVID-19 vaccination date (N=172 316)** [a]

| Outcome | BMI Status | Incident/total COVID (%) [b] | RR | LCI | UCI |
|---|---|---|---|---|---|
| PASC (U09.9) | Healthy weight | 514/68 918 (0.7) | 1 [Ref] | 1 [Ref] | 1 [Ref] |
| | Overweight | 137/17 605 (0.8) | 1.046 | 0.867 | 1.262 |
| | Obesity | 199/25 372 (0.8) | **1.251** | **1.062** | **1.475** |
| | Severe obesity | 552/60 241 (0.9) | **1.422** | **1.254** | **1.611** |
| | *P* for trend | NA | | 0.001 | |
| PASC symptoms and conditions | **Any Occurrences** | | | | |
| | Healthy weight | 28 674/68 918 (41.6) | 1 [Ref] | 1 [Ref] | 1 [Ref] |
| | Overweight | 7 637/17 605 (43.4) | 1.029 | 0.982 | 1.079 |
| | Obesity | 11 081/25 372 (43.7) | **1.106** | **1.062** | **1.152** |
| | Severe obesity | 26 925/60 241 (44.7) | **1.175** | **1.138** | **1.213** |
| | *P* for trend | NA | | <0.001 | |
| | **Total Occurrences** | | | | |
| | Healthy weight | NA | 1 [Ref] | 1 [Ref] | 1 [Ref] |
| | Overweight | NA | 1.052 | 0.999 | 1.108 |
| | Obesity | NA | **1.135** | **1.086** | **1.186** |
| | Severe obesity | NA | **1.182** | **1.142** | **1.223** |
| | *P* for trend | NA | | <0.001 | |

Abbreviation: BMI, body mass index; LCI, lower 95% confidence interval; NA, not applicable; PASC, post-acute sequelae of SARS-CoV-2 infection; RR, relative risk; UCI, upper 95% confidence interval

[a] Adjusted for age assessed BMI and entered cohort (continuous), sex, race/ethnicity, PMCA index, predominant variant, acute COVID-19 severity, numbers of emergency department visits, outpatient department visits, inpatient department visits, medications or prescriptions, negative COVID-19 tests, doses of COVID-19 vaccine before infection and interval since last COVID-19 vaccination date.

[b] Incident referred to the count of participants developed the outcome we interested in, total COVID referred to the count of the participants in the corresponding group, and the value in the bracket referred to the percentage of the groups who developed the outcome we interested in.

**eTable 11. Estimated association of BMI status prior to the SARS-CoV-2 infection and risk of PASC adjusting type of insurance (N=140 036) [a]**

| Outcome | BMI Status | Incident/total COVID (%) [b] | RR | LCI | UCI |
|---|---|---|---|---|---|
| PASC (U09.9) | Healthy weight | 396/54 114 (0.7) | 1 [Ref] | 1 [Ref] | 1 [Ref] |
| | Overweight | 117/14 233 (0.8) | 1.146 | 0.932 | 1.410 |
| | Obesity | 166/21 047 (0.8) | **1.360** | **1.131** | **1.635** |
| | Severe obesity | 460/50 642 (0.9) | **1.547** | **1.315** | **1.819** |
| | *P* for trend | NA | | 0.002 | |
| PASC symptoms and conditions | **Any Occurrences** | | | | |
| | Healthy weight | 22 624/54 114 (41.8) | 1 [Ref] | 1 [Ref] | 1 [Ref] |
| | Overweight | 6 197/14 233 (43.5) | 1.018 | 0.965 | 1.075 |
| | Obesity | 9 037/21 047 (42.9) | **1.056** | **1.008** | **1.106** |
| | Severe obesity | 22 482/50 642 (44.4) | **1.064** | **1.021** | **1.109** |
| | *P* for trend | NA | | <0.001 | |
| | **Total Occurrences** | | | | |
| | Healthy weight | NA | 1 [Ref] | 1 [Ref] | 1 [Ref] |
| | Overweight | NA | 1.031 | 0.972 | 1.094 |
| | Obesity | NA | **1.072** | **1.019** | **1.127** |
| | Severe obesity | NA | **1.066** | **1.019** | **1.115** |
| | *P* for trend | NA | | <0.001 | |

Abbreviation: BMI, body mass index; LCI, lower 95% confidence interval; NA, not applicable; PASC, post-acute sequelae of SARS-CoV-2 infection; RR, relative risk; UCI, upper 95% confidence interval

[a] Adjusted for age assessed BMI and entered cohort (continuous), sex, race/ethnicity, PMCA index, predominant variant, acute COVID-19 severity, numbers of emergency department visits, outpatient department visits, inpatient department visits, medications or prescriptions, negative COVID-19 tests, and type of insurance.

[b] Incident referred to the count of participants developed the outcome we interested in, total COVID referred to the count of the participants in the corresponding group, and the value in the bracket referred to the percentage of the groups who developed the outcome we interested in.

**eTable 12. Estimated association of BMI status prior to the SARS-CoV-2 infection and risk of PASC after excluding participants with diabetes (N=169 495) [a]**

| Outcome | BMI Status | Incident/total COVID (%) [b] | RR | LCI | UCI |
|---|---|---|---|---|---|
| PASC (U09.9) | Healthy weight | 508/67 978 (0.7) | 1 [Ref] | 1 [Ref] | 1 [Ref] |
| | Overweight | 136/17 244 (0.8) | 1.058 | 0.876 | 1.276 |
| | Obesity | 196/24 951 (0.8) | **1.258** | **1.066** | **1.484** |
| | Severe obesity | 548/59 322 (0.9) | **1.433** | **1.263** | **1.626** |
| | *P* for trend | NA | | 0.001 | |
| PASC symptoms and conditions | **Any Occurrences** | | | | |
| | Healthy weight | 28 302/68 918 (41.6) | 1 [Ref] | 1 [Ref] | 1 [Ref] |
| | Overweight | 7 487/17 605 (43.4) | 1.032 | 0.984 | 1.083 |
| | Obesity | 11 894/25 372 (43.7) | **1.111** | **1.067** | **1.157** |
| | Severe obesity | 26 486/60 241 (44.6) | **1.178** | **1.141** | **1.217** |
| | *P* for trend | NA | | <0.001 | |
| | **Total Occurrences** | | | | |
| | Healthy weight | NA | 1 [Ref] | 1 [Ref] | 1 [Ref] |
| | Overweight | NA | 1.052 | 0.999 | 1.108 |
| | Obesity | NA | **1.138** | **1.089** | **1.189** |
| | Severe obesity | NA | **1.186** | **1.146** | **1.228** |
| | *P* for trend | NA | | <0.001 | |

Abbreviation: BMI, body mass index; LCI, lower 95% confidence interval; NA, not applicable; PASC, post-acute sequelae of SARS-CoV-2 infection; RR, relative risk; UCI, upper 95% confidence interval

[a] Adjusted for age assessed BMI and entered cohort (continuous), sex, race/ethnicity, PMCA index, predominant variant, acute COVID-19 severity, numbers of emergency department visits, outpatient department visits, inpatient department visits, medications or prescriptions, and negative COVID-19 tests

[b] Incident referred to the count of participants developed the outcome we interested in, total COVID referred to the count of the participants in the corresponding group, and the value in the bracket referred to the percentage of the groups who developed the outcome we interested in.

**eTable 13. Estimated association of BMI status prior to the SARS-CoV-2 infection and risk of PASC after excluding obese participants taking weight-loss drugs (N=169 255) [ab]**

| Outcome | BMI Status | Incident/total COVID (%) [c] | RR | LCI | UCI |
|---|---|---|---|---|---|
| PASC (U09.9) | Healthy weight | 514/68 918 (0.7) | 1 [Ref] | 1 [Ref] | 1 [Ref] |
| | Overweight | 137/17 605 (0.8) | 1.046 | 0.867 | 1.261 |
| | Obesity | 189/24 432 (0.8) | **1.256** | **1.062** | **1.485** |
| | Severe obesity | 531/58 300 (0.9) | **1.429** | **1.259** | **1.623** |
| | *P* for trend | NA | | 0.002 | |
| PASC symptoms and conditions | **Any Occurrences** | | | | |
| | Healthy weight | 28 674/68 918 (41.6) | 1 [Ref] | 1 [Ref] | 1 [Ref] |
| | Overweight | 7 637/17 605 (43.4) | 1.030 | 0.982 | 1.080 |
| | Obesity | 10 543/25 372 (43.2) | **1.115** | **1.070** | **1.161** |
| | Severe obesity | 25 800/60 241 (44.3) | **1.184** | **1.146** | **1.222** |
| | *P* for trend | NA | | <0.001 | |
| | **Total Occurrences** | | | | |
| | Healthy weight | NA | 1 [Ref] | 1 [Ref] | 1 [Ref] |
| | Overweight | NA | **1.053** | **1.00** | **1.109** |
| | Obesity | NA | **1.140** | **1.091** | **1.192** |
| | Severe obesity | NA | **1.192** | **1.151** | **1.234** |
| | *P* for trend | NA | | <0.001 | |

Abbreviation: BMI, body mass index; LCI, lower 95% confidence interval; NA, not applicable; PASC, post-acute sequelae of SARS-CoV-2 infection; RR, relative risk; UCI, upper 95% confidence interval

[a] Weight-loss drugs included Metformin, Orlistat, Liraglutide, Exenatide, Dulaglutide, Semaglutide, Setmelanotide, Phentermine Topiramate

[b] Adjusted for age assessed BMI and entered cohort (continuous), sex, race/ethnicity, PMCA index, predominant variant, acute COVID-19 severity, numbers of emergency department visits, outpatient department visits, inpatient department visits, medications or prescriptions, and negative COVID-19 tests

[c] Incident referred to the count of participants developed the outcome we interested in, total COVID referred to the count of the participants in the corresponding group, and the value in the bracket referred to the percentage of the groups who developed the outcome we interested in.

**eTable 14. Estimated association of BMI status prior to the SARS-CoV-2 infection and risk of PASC based on primary care sites (N=42 470) [a]**

| Outcome | BMI Status | Incident/total COVID (%) [b] | RR | LCI | UCI |
|---|---|---|---|---|---|
| PASC (U09.9) | Healthy weight | 172/25 890 (0.7) | 1 [Ref] | 1 [Ref] | 1 [Ref] |
| | Overweight | 54/7 083 (0.8) | 1.175 | 0.869 | 1.590 |
| | Obesity | 63/7 674 (0.8) | **1.450** | **1.082** | **1.944** |
| | Severe obesity | 18/1 823 (1.0) | 1.137 | 0.699 | 1.848 |
| | *P* for trend | NA | | 0.043 | |
| PASC symptoms and conditions | **Any Occurrences** | | | | |
| | Healthy weight | 9 335/25 890 (36.1) | 1 [Ref] | 1 [Ref] | 1 [Ref] |
| | Overweight | 2 684/7 083 (37.9) | 1.057 | 0.978 | 1.141 |
| | Obesity | 3 010/7 674 (39.2) | **1.080** | **1.002** | **1.164** |
| | Severe obesity | 817/1 823 (44.8) | **1.106** | **0.942** | **1.299** |
| | *P* for trend | NA | | <0.001 | |
| | **Total Occurrences** | | | | |
| | Healthy weight | NA | 1 [Ref] | 1 [Ref] | 1 [Ref] |
| | Overweight | NA | **1.102** | **1.013** | **1.198** |
| | Obesity | NA | **1.118** | **1.029** | **1.213** |
| | Severe obesity | NA | 1.169 | 0.981 | 1.392 |
| | *P* for trend | NA | | <0.001 | |

Abbreviation: BMI, body mass index; LCI, lower 95% confidence interval; NA, not applicable; PASC, post-acute sequelae of SARS-CoV-2 infection; RR, relative risk; UCI, upper 95% confidence interval

[a] Adjusted for age assessed BMI and entered cohort (continuous), sex, race/ethnicity, PMCA index, predominant variant, acute COVID-19 severity, numbers of emergency department visits, outpatient department visits, inpatient department visits, medications or prescriptions, and negative COVID-19 tests

[b] Incident referred to the count of participants developed the outcome we interested in, total COVID referred to the count of the participants in the corresponding group, and the value in the bracket referred to the percentage of the groups who developed the outcome we interested in.

**eTable 15. Estimated association of BMI status prior to the SARS-CoV-2 infection and risk of foreign body in ear as a negative control outcome (N=172 316) [a]**

| BMI Status | Incident/total COVID (%) [b] | RR | LCI | UCI |
|---|---|---|---|---|
| Healthy weight | 6/68 918 (0.0) | 1 [Ref] | 1 [Ref] | 1 [Ref] |
| Overweight | 4/17 605 (0.0) | 2.617 | 0.705 | 9.706 |
| Obesity | 3/25 372 (0.0) | 0.942 | 0.245 | 3.622 |
| Severe obesity | 5/60 241 (0.0) | 0.790 | 0.246 | 2.536 |
| *P* for trend | NA | | 0.852 | |

Abbreviation: BMI, body mass index; LCI, lower 95% confidence interval; NA, not applicable; RR, relative risk; UCI, upper 95% confidence interval

[a] Adjusted for age assessed BMI and entered cohort (continuous), sex, race/ethnicity, PMCA index, predominant variant, acute COVID-19 severity, numbers of emergency department visits, outpatient department visits, inpatient department visits, medications or prescriptions, and negative COVID-19 tests

[b] Incident referred to the count of participants developed the outcome we interested in, total COVID referred to the count of the participants in the corresponding group, and the value in the bracket referred to the percentage of the groups who developed the outcome we interested in.
